# EBV-positive Hodgkin lymphoma with immunosuppressive microenvironment during IL-23 inhibitor therapy for psoriasis

**DOI:** 10.1101/2025.04.08.25325422

**Authors:** Bhaumik Shah, Janusz Franco-Barraza, Kathy Q. Cai, Mariia Dmitrieva, Marcus Messmer, Yibin Yang, Mariusz Wasik, Edna Cukierman, Reza Nejati

**Affiliations:** Department of Pathology, Cancer Signaling & Microenvironment Program, Philadelphia, Pennsylvania, USA; Department of Pathology, Fox Chase Cancer Center, Temple Health, Philadelphia, Pennsylvania, USA

## Abstract

We report the initial case of EBV-positive classic Hodgkin lymphoma (cHL) after guselkumab therapy for psoriasis. High-plex spatial proteomic analysis showed a Th2-polarized immunosuppressive tumor microenvironment, indicating immune reprogramming from IL-23 inhibition might have triggered the onset of EBV-positive cHL.

## Introduction

In the DISCOVER extension studies, guselkumab use demonstrated a favorable safety profile without any incidence of lymphoma in biologic-naive patients with active psoriatic arthritis (PsA)^1,2^. We report the first case of Epstein-Barr virus (EBV)-positive classic Hodgkin lymphoma (cHL) occurring in a biologic-naive PsA patient during guselkumab (IL-23 inhibitor) therapy. Spatial proteomic analysis of the immune tumor microenvironment (TME), using single-cell high-plex sequential cycling immunofluorescence (seqIF™)^3^, revealed an immunosuppressive profile enriched in T helper 2 (Th2) cells, contrasting with the typical T helper 1 (Th1) profile of EBV-positive cHL^4,5^. These findings suggest IL-23 inhibition might have reprogrammed the immune response, promoting the emergence of EBV-positive cHL.

### Case Summary

A young adult male with a 10-year history of chronic plaque psoriasis was placed on guselkumab (IL-23 inhibitor) therapy for active PsA. Three to four months later, he experienced night sweats and recurrent head-turning syncope episodes. Approximately seven months into therapy, a painless, non-mobile right cervical lump was discovered. Contrast-enhanced computed tomography (CECT) of the neck revealed bulky right cervical lymphadenopathy, and an 18F-FDG PET/CT scan (skull vertex to thigh) detected a large hypermetabolic lymph node conglomerate (8.8 cm) in the right cervical region, involving lymph node group levels I to V (Fig. 1A-C). Right level III lymph nodes (highest SUV 18.8) were excised (Fig.1 C). Excisional biopsy revealed partially preserved lymph node architecture with scattered Hodgkin and Reed-Sternberg (HRS) cells, predominantly in the interfollicular area (Fig. 1D). Immunohistochemistry confirmed the characteristic HRS cell phenotype (Fig. 1E-G). HRS cells were strongly positive for Epstein-Barr encoding region (EBER) in situ hybridization (Fig. 1H) and EBV latent membrane protein (LMP1; Fig. 1I), while negative for EBV nuclear antigen-2 (EBNA-2), indicating a latency pattern IIa (LMP-1+, EBNA-2-). Plasma EBV DNA level (quantitative RT-PCR) detected a viral load of 75 IU/ml (1.88 Log10 IU/ml) within a quantification range of 35.0 to 100,000,000 IU/mL (1.54 to 8.00 Log10 IU/mL). Chromosomal microarray analysis yielded negative results, but next generation sequencing (275-gene panel) detected a Tier 2 splice site variant in the *ATR* gene (c.3819+1G>T) with a variant allele frequency (VAF) of 57.1%, indicating a potentially germline variant (Table J). The diagnosis of EBV-positive cHL, possibly iatrogenic or related to immunomodulator (IL-23 inhibitor) therapy, was made. Guselkumab was discontinued. However, two to three weeks later, due to the large mass and syncope episodes, the patient underwent two cycles of ABVD regimen followed by 20 Gy consolidation ISRT (10 fractions). Follow-up FDG PET/CT showed a complete metabolic response (Deauville score 2). Six months post-diagnosis, he was started on apremilast, a small-molecule phosphodiesterase 4 (PDE4) inhibitor for psoriatic arthritis. At the time of this report, the patient remains in remission with no complaints after six months of apremilast therapy.

**Figure 1.**
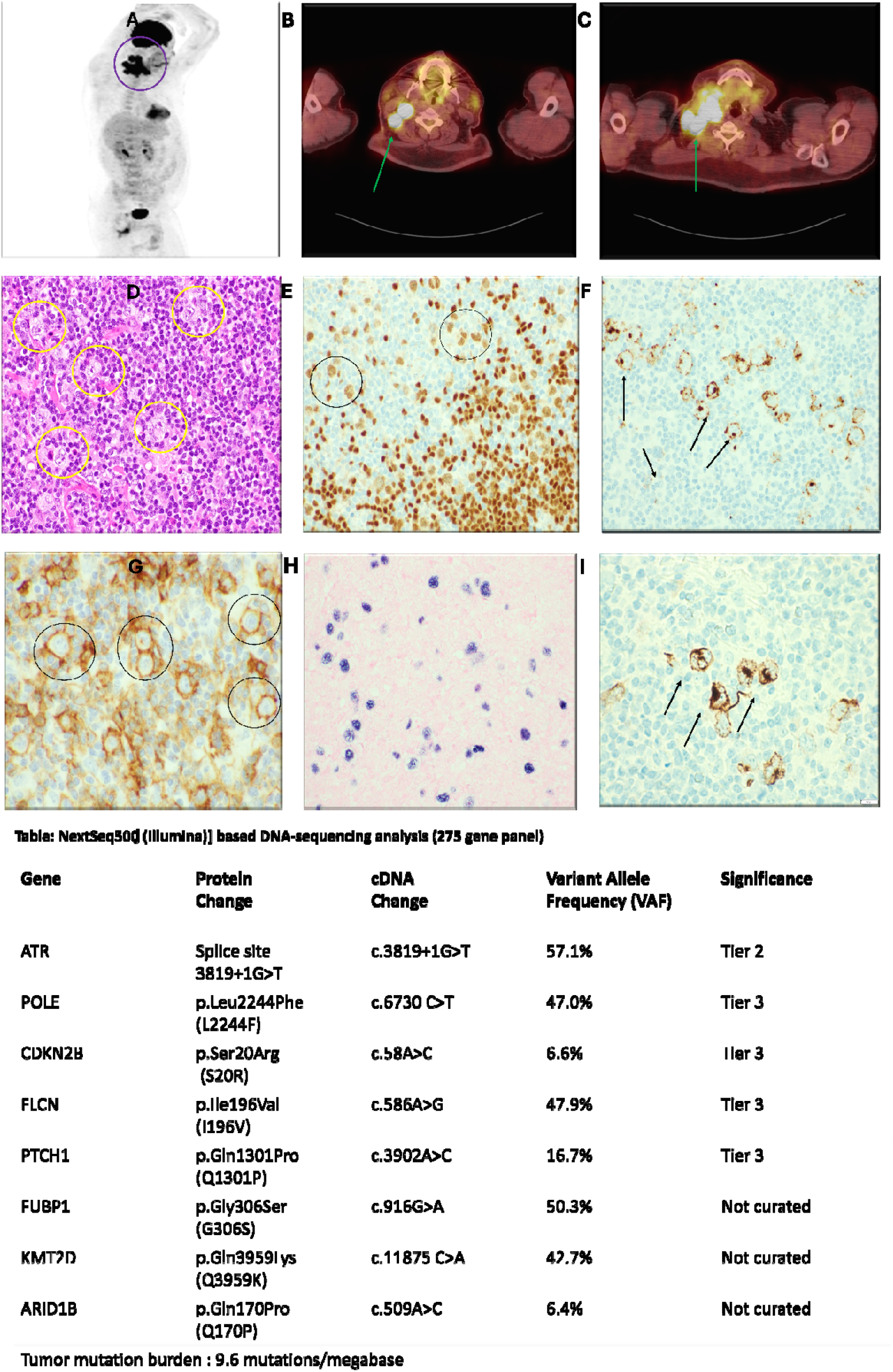
EBV-positive classic Hodgkin lymphoma following guselkumab (IL-23 inhibitor) therapy. (**A)** Maximum intensity projection sagittal FDG PET image shows an FDG-avid, large (8.8 cm), right cervical lymph node conglomerate (purple circle) involving Level I to V lymph node groups. **(B, C)** Fused transaxial FDG PET/CT image shows intensely increased FDG uptake (long green arrow) in the right cervical level II (Panel B) & level III (maximum SUV 18.8; Panel C) lymph node group. **(D)** Representative H&E-stained-section (40x objective) of the right cervical lymph node shows scattered large mononuclear and binucleated/multinucleated atypical cells with prominent nucleoli, consistent with HRS cells (yellow circle), predominantly in the interfollicular area (follicles are not shown for comparison) (**E-H)** Immunohistochemical staining of representative sections from excised right level III lymph nodes. PAX5 shows weak to moderate staining (black circle) in HRS cells (Panel E; 10x objective); CD30 shows strong membranous and Golgi patterns (long black arrow) in HRS cells (Panel F; 40x objective); Strong CD200-positivity^6^ (black circle) in HRS cells helps differentiate EBV-positive cHL from EBV-positive diffuse large B-cell lymphoma, which shows negative to partial/weak staining (Panel G; 40x objective). EBER by in situ hybridization is positive in HRS cells (Panel H; 40x objective). **(I**) LMP1 is strongly expressed in HRS cells (Panel I; 40x objective). **(J)** The results of NGS [NextSeq500 (Illumina)] based DNA-sequencing analysis (275 gene panel). *ATR, CDKN2B, PTCH1, KMT2D, ARID1B, POLE, FLCN, FUB1* genes may be implicated in the epigenetic regulation of EBV latency and EBV lytic reactivation^7,8^. The clinical significance of the detected variants in epigenetic regulation of EBV latency is not known. Abbreviations: EBV, Epstein-Barr virus; FDG PET, fluorodeoxyglucose positron emission tomography; CT, computed tomography; SUV max, maximum standardized uptake value; H&E, Hematoxylin and eosin stain; HRS cells, Hodgkin and Reed-Sternberg cells; EBER, Epstein-Barr encoding region; ISH, in situ hybridization; *ATR* ataxia telangiectasia and Rad3 related; *POLE*, DNA polymerase epsilon, catalytic subunit; CDKN2B, cyclin dependent kinase inhibitor 2B; FLCN, Folliculin; PTCH1, Patched 1, FUBP1, far upstream element binding protein 1; KMT2D: Lysine methyltransferase 2D; ARID1B, AT-rich interaction domain 1B.

### High-plex spatial proteomic analysis

We analyzed the TME to investigate guselkumab’ s immunomodulatory effects and its potential link to EBV-positive cHL. Unstained slides from FFPE sections of leftover diagnostic LN were analyzed using the Lunaphore’s COMET™ protocol^3^ designed for spatial single-cell high-plex seqIF™ (Fig. 2). A customized biomarker signature was used to characterize the cHL TME. Nine markers (CD20, T-bet, CD4, CD163, CCR4, CCR6, HLA-DR, Ki67, CTLA4) were spatially mapped to detect HRS cells, and determine individual immune cell composition, utilizing HORIZON™ software, which integrates single-cell segmentation and rule-based classification algorithms. Ten cell populations were identified, including HRS cells, B cell subsets, Th1 cells, Th2 cells, Th17 cells, immunosuppressive T-cells, and macrophages (Fig. 2F-H). Single-cell proteomics revealed an immunosuppressive TME, with predominant Th2 cells (CD20^-^, CD4^+^, T-bet^-^, CCR6^-^, CCR4^+^, HLA-DR^+^), numerous immunosuppressive T-cells (CD20-, CD4+, T-bet-, CCR4+, CTLA4+), a low Th1/Th2 ratio (0.10), and reduced Th17 cells (CD20^-^, CD4^+^, T-bet^-^, CCR6^+^, CCR4^-^, HLA-DR^+ or -^) (Fig 2). Contrasting with the typical Th1 profile of EBV-positive cHL, our TME study showed a Th2-dominant profile, suggesting immune milieu reprogramming as a possible driver of EBV-positive cHL emergence.

**Figure 2.**
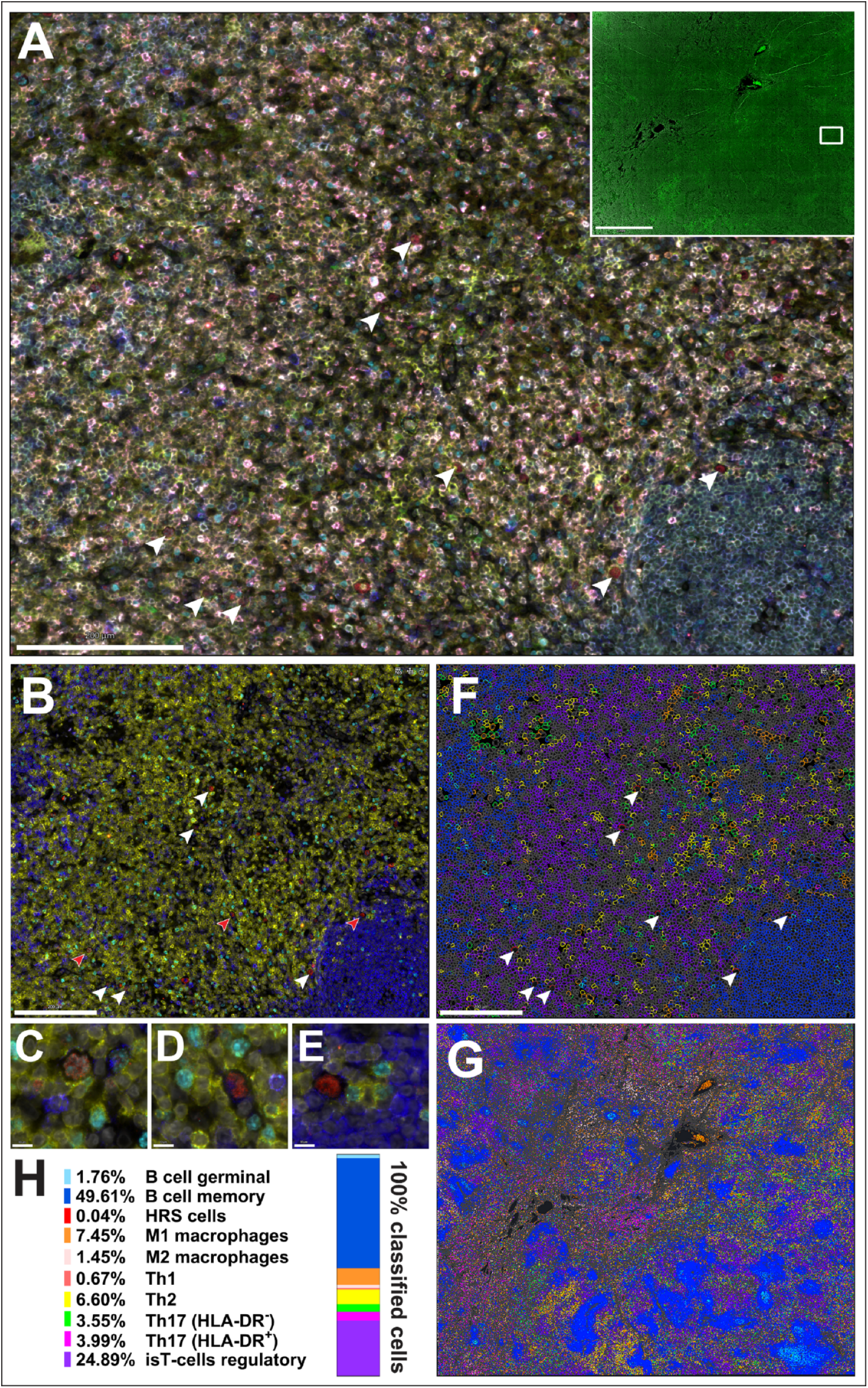
Spatial single-cell proteomic seqIF analysis reveals an immunosuppressive lymph node TME. (A-E) Representative high-plex seqIF^3^ images showing 10 distinct cell populations (B cells, memory or germinal; HRS; M1 macrophages; M2 macrophages; Th1 cells; Th2 cells; Th17 cells, HLA-DR neg or pos; and immunosuppressive (is)-T cells [isT-cells]) using nine markers (CD20: blue, CD4: yellow, CD163: magenta, CCR6: green, CCR4: light gray, HLA-DR: orange, CTLA-4: purple, T-bet: red, and Ki67: cyan). The cell population is defined as follows: B germinal center (CD20^+^, HLA-DR^+^, CCR6^+^, Ki67^+^); B memory (CD20^+^, HLA-DR^+^, CCR6^+^, Ki67^-^); HRS (CD20^-^/low, T-bet^+^); Macrophages M1 (CD20^-^, CD4^-^, HLA-DR^+^); M2 (CD20^-^, CD4^-^, CD163^+^); Th1 (CD20^-^, CD4^+^, T-bet^+^); Th2 (CD20^-^, CD4^+^, T-bet^-^, CCR6^-^, CCR4^+^, HLA-DR^+^); Th17 (CD20^-^, CD4^+^, T-bet^-^, CCR6^+^, CCR4^-^, HLA-DR^+ or -^); immunosuppressive (is)-T-cells (CD20^-^, CD4^+^, T-bet^-^, CCR4^+^, CTLA4^+^) (A) Composite marker visualization highlights immune cellular heterogeneity. Insert shows the entire tissue section analyzed, visualized by the tissue’s autofluorescence (green) and cellular nuclei (gray); the white rectangle marks the zoomed-in region shown in B-F. (B) Proliferating (Ki67+) and non-proliferating B cells (CD20+), T cells (CD4+), and HRS cells (large nucleus, CD20?/low, T-bet+) are shown, with HRS cells (white/red arrows, also marked in F, all white) distributed within interfollicular areas and interacting with T cells. (C-E) High-magnification views of HRS cells (from red arrows in B) show T-bet+ enlarged nuclei, low/no CD20, and no CD4 expression. (F) Example of HORIZON-driven single cell detection and classification of populations (from B), rendering color-coded polygons based on phenotype-specific marker combinations. (G) Spatial mapping of assorted classified immune cell populations (colored dots as shown in H) across the lymph node tissue (unclassified cells appear in dark gray). (H). Proportions of classified cell populations are shown in a color-coded percentage graph. Scale bars: A, B, and F = 200 µm; insert in A = 2 mm, C-E = 10 µm.

## Discussion

Psoriasis, driven by IL-23/IL-17 axis hyperactivation, is an independent risk factor for lymphoma, particularly cHL^9^. IL-23 inhibitors (guselkumab, tildrakizumab, risankizumab, mirikizumab) and IL-17 inhibitors (secukinumab, Ixekizumab, bimekizumab, and brodalumab) are commonly used to treat immune-mediated inflammatory diseases, particularly moderate-to-severe psoriasis (PsO), PsA and inflammatory bowel disease^10^. IL-23 inhibitors target IL-23, a key upstream IL-12 family cytokine driving Th17 cell differentiation via JAK2/STAT3 phosphorylation, while IL-17 inhibitors block IL-17, a major effector of chronic inflammation and autoimmunity, downstream of IL-23^11^. Guselkumab, a fully human IgG1λ monoclonal antibody, a selective IL23p19 subunit-inhibitor, has demonstrated a favorable safety profile, in an integrated analysis of eleven clinical trials (n=4,399 PsO/PsA patients, 5-year follow-up) with no increased lymphoma risk^12,13^. However, two unspecified lymphoma cases were reported in the VOYAGE extension studies, involving 1721 guselkumab-treated moderate-to-severe PsO patients^14,15^. Meta-analysis^16^ and a global cohort study^17^ also reported no increased risk of malignancy with IL-17/IL-23 inhibitors and reported highest drug survival rates^18^ for guselkumab and risankizumab. A post-marketing surveillance study reported one non-lymphoma malignancy amongst guselkumab-treated PsO patients (n=411)^19,20^.

Only one prior case of cHL has been documented following IL-23/IL-12 inhibitor (ustekinumab) therapy in a young adult female with PsO. Like our case, cHL was diagnosed after just two months of treatment. Both patients had a 10-year history of chronic plaque psoriasis without prior malignancy. While PsO itself increases cHL risk, potentially via NF-kB and JAK2/STAT signaling, the short interval between treatment initiation and cHL diagnosis contrasts with the typically long latency period for lymphomagenesis. The EBV and TME status of the previously reported case remains unknown. However, in our case, EBV, a known driver of lymphoma and autoimmune diseases ^21^, demonstrated a restricted latency IIa viral gene expression program in HRS cells along with a low viral load-a marker of EBV activity. While EBV-positive cHL usually exhibits a Th1-dominant profile, and EBV-negative cHL a Th17 profile, this case displayed CCR4+ CCR6-Th2 dominance, a scarcity of Th1 cells, and decreased Th17 cells, suggesting Th17 cell plasticity and Th2 polarization under IL-23/IL-17 blockade. Latency IIa-expressing germinal center B-cells (LMP1+, EBNA1+, EBNA2-) are considered precursors of EBV-associated cHL^22^. Both LMP and EBNA1 promote Treg recruitment, further enhanced by Th2 polarization^23^. EBV hijacks host epigenetic mechanisms to alter DNA methylation patterns, histone modifications and chromatin remodeling in both viral and host genomes to regulate its latency and reactivation^7^. Although, the role of the detected variants in *ATR, CDKN2B, PTCH1, KMT2D, ARID1B, POLE, FLCN, FUB1* genes (Fig 1; Panel J) is not clear, alterations in these DNA repair genes (*ATR, POLE*), histone methyltransferase (*KMT2D*), chromatin remodeler (*ARID1B*), tumor suppressor genes (*CDKN2B, FLCN, PTCH1*) may be implicated in epigenetic dysregulation, influencing the switch in EBV latency program^7,8^. IL-23 blockade may have fostered a favorable cytokine milieu for EBV to shift its latency program via epigenetic regulators^24^, potentially facilitating cHL emergence via LMP1-induced NF-kB dysregulation.

## Conclusion

These findings indicate that guselkumab may trigger immune reprogramming toward an immunosuppressive Th2 profile. This may have occurred in this case, despite the drug’s consistent favorable safety profile observed across multiple clinical trials and extension studies. The short interval between exposure and cHL diagnosis in these two patients justifies ongoing post-marketing surveillance to evaluate the lymphoma risk and incidence, particularly in EBV-positive PsO and PsA patients. Pre-screening PsO patients for EBV prior to initiating IL-23 inhibitors is, therefore, recommended to identify at-risk individuals. Larger studies are required to identify predictive markers and stratify patients unsuitable for newer biologic therapies.

## Data Availability

All data produced in the present study are available upon reasonable request to the authors

## Acknowledgments

The biospecimens and clinical data being utilized in this tumor microenvironment study were collected previously as part of the patient’s routine clinical care. No separate specimens were collected for the study. The Institutional Review Board of Fox Chase Cancer Center provided ethical approval for this work. All authors state that the presented material does not include any information, unique characteristics or identifiers that could be used alone or in combination with other information to identify, directly or indirectly, an individual who is a subject of the case report. Funds supporting this study were from NCI/NIH R01 CA269660-01 (EC and JFB), U54 CA272686 (EC) and the comprehensive cancer center core grant P30 CA006927 in support of the histopathology (housing the spatial high-plex proteomic seqIF and digital analysis branch) facilities at the Fox Chase Cancer center. In addition, funds supporting this study were obtained from the ACS grant RP-23-1070169-01-WRP (EC), The trustees of the Community Foundation of New Jersey on behalf of Mrs. Marina P Zazanis (EC), The Pancreatic Cure Foundation (EC, JFB, MD), and the 5th AHEPA Cancer Research Foundation, Inc. (EC). The preliminary findings from this study were presented by B.S. before the expert panel as part of the oral presentation, titled “Classic Hodgkin lymphoma, EBV+, following immunomodulator therapy (IL-23 inhibitor) for Psoriasis” at the 22nd Meeting of the European Association for Hematopathology, held in Dubrovnik, Croatia, Sep 21-Sep 26, 2024. The authors thank Dr. J. Kowal and E. Geneletti, current or former Lunaphore employees, for their expertise and support in developing the high-plex immunofluorescence analysis using HORIZON™ image analysis software from the COMET™ suite.

## Authorship

Original idea: R.N.

Diagnostic work up: B.S. (lead), R.N. (equal).

Conceptualization, review of literature and hypothesis: B.S. (lead), R.N. (equal), J.F. (supporting), E.C. (supporting)

Methodology (design, optimization & validation of the antibodies): J.F (lead), K.Q. (equal), B.S. (supporting), R.N. (supporting), E.C. (supporting).

Data acquisition (performance of the experiment) and data validation: J.F. (lead), K.Q. (equal);

B.S. (supporting; validation)

Data curation and data analysis: J.F. (lead), M.D. (equal), B.S. (supporting; strategy) Data interpretation: B.S. (lead), J.F. (equal), R.N (supporting), E.C. (supporting), M.W. (supporting).

Writing original draft: B.S (lead), J.F(lead; material and methods) Data visualization: B.S. (lead; figure 1), J.F (lead; figure 2)

Critical review and editing of the manuscript: B.S. (lead), E.C (equal), J.F. (equal), R.N. (equal),

M.W. (equal), M.M. (supporting), Y.Y. (supporting).

Project supervision and administration: R.N. (lead), B.S(equal), J.F(equal), E.C(equal), M.W. (equal)

Resources and Funding acquisition: E.C (lead), R.N (equal), M.W. (equal), Y.Y (supporting).

## Data availability

Detailed materials and methods, including spatial single-cell high-plex seqIF™ staining protocol, methodology for data preprocessing, filtering and analysis (cell segmentation, supervised and unsupervised phenotyping and spatial analysis) and the original OME-TIFF files containing the multiplex immunofluorescence data underlying the findings described in this manuscript will be made available upon reasonable request to the corresponding author.

